# Characterising the motif composition and allele length distribution of *ZFHX3* GGC repeat expansions in amyotrophic lateral sclerosis

**DOI:** 10.64898/2026.03.09.26347973

**Authors:** Zoe N. Zussa, Andrew N. Smith, Joke J.F.A van Vugt, Daniel S. O’Shaughnessy, Natalie Grima, Sandrine Chan Moi Fat, Ian P. Blair, Dominic B Rowe, Roger Pamphlett, Garth A. Nicholson, Matthew C.K Kiernan, Wouter van Rheenen, Jan Veldink, Project MinE ALS sequencing consortium, Kelly L. Williams, Lyndal Henden

## Abstract

**Background and objectives:** A pathogenic GGC repeat expansion in the zinc finger homeobox 3 (*ZFHX3*) gene, encoding a pure polyglycine tract, is the cause of spinocerebellar ataxia type 4 (SCA4). Intermediate expansions of other SCA loci contribute to the risk of amyotrophic lateral sclerosis (ALS), a fatal neurodegenerative disease involving the progressive loss of motor neurons. There is increasing awareness of the role of short tandem repeat (STR) motif composition and configuration in disease pathogenicity. Given the genetic pleiotropy between ALS and SCA, this study aimed to evaluate whether *ZFHX3* GGC expansions were associated with ALS and to characterise repeat motif composition.

**Methods:** ExpansionHunter v5 was used to genotype *ZFHX3* GGC repeat sizes in short-read whole genome sequencing data from people with ALS and healthy controls of European ancestry. Repeat sizes were visually inspected using REViewer v2. Repeat motif configurations of Australian ALS cases and healthy controls were manually derived from REViewer images. Receiver operating characteristic (ROC) curve analysis and Youden’s J statistic were performed to find a candidate repeat size threshold for association testing. Fisher’s exact tests were performed to evaluate the associations of repeat size and motif composition with disease status.

**Results:** Analysis of 5,785 people with ALS and 7,982 healthy controls found no association between *ZFHX3* GGC repeat expansions and disease risk. Fifty unique repeat motif compositions were identified across 802 people with ALS and 800 healthy controls. Of these, eleven distinct configurations coded a pure polyglycine tract which, when expanded, is canonical to SCA4, though no association with ALS was found.

**Discussion:** Although no association was observed between *ZFHX3* GGC repeat expansions and ALS, this study established the dynamic nature of *ZFHX3* repeat motif composition and configuration. Unique motif compositions were identified both within and between repeat sizes, including the presence of pure polyglycine repeats. Consideration of repeat motif composition and configuration, in addition to repeat allele length, may be important for assessing neurodegenerative disease risk.

## Introduction

Amyotrophic lateral sclerosis (ALS) is a fatal neurodegenerative disease characterised by progressive degeneration of upper and lower motor neurons, resulting in muscle weakness, paralysis and death, typically due to respiratory failure within 3-5 years of symptom onset^1^. Gene mutations are the only proven cause of ALS, accounting for two-thirds of familial (inherited) ALS cases and ∼20% of sporadic ALS cases^2^. ALS appears genetically heterogenous, with more than 30 genes and over 1,000 genetic variants implicated in disease risk or causality to date^3^. The most common genetic cause of ALS is a pathogenic short tandem repeat (STR) expansion in the intron of the *C9orf72* gene which accounts for disease in 40% of familial and 10% of sporadic cases^1^. The genetic landscape of ALS is further complicated by pleiotropy where the same mutation can give rise to different phenotypes and different disease manifestations. Notably the *C9orf72* repeat expansion is also the most common genetic cause of frontotemporal dementia (FTD); a neurodegenerative disease characterised by behavioural and cognitive decline^4^. Pleiotropy in ALS extends beyond *C9orf72*, with increasing genetic evidence linking ALS and spinocerebellar ataxia (SCA); a group of neurodegenerative disorders marked by progressive loss of balance, coordination and slurred speech^5^. Pathogenic CAG repeat expansions in *ATXN1* and *ATXN2* cause SCA1 and SCA2, respectively, while intermediate-length CAG expansions in these genes have been associated with increased ALS risk^6,7,8^.

Recently, a coding pathogenic GGC repeat expansion in exon 10 of the zinc finger homeobox 3 gene (*ZFHX3*) was identified as the cause of SCA4 in families with Northern European ancestry, placing SCA4 among the growing group of polyglycine (polyG) expansion disorders^9,10,11^. *ZFHX3* encodes a transcription factor for neuronal and myogenic cell differentiation and cell proliferation^12^. Pure polyG repeat expansions in *ZFHX3* exceeding 41 GGC repeats lead to a pathogenic gain of gene function resulting in neuron degeneration, which is a hallmark pathological feature of both SCA and ALS^9^. Interestingly, individuals with SCA4 also show loss of motor neurons in the anterior horns and degradation of posterior tracts of the spinal cord, consistent with ALS pathology^10^.

Investigations of STRs in polyG disorders and SCAs have emphasised the importance of motif composition (i.e. the exact sequence of nucleotides or amino acids in the expanded repeat) given that the presence and configuration of synonymous or missense interrupting motifs can modify disease phenotype and clinical presentation^13,14,15^. Synonymous interruptions are nucleotide changes that alter repeat motif composition without changing the encoded amino acid and have been demonstrated to occur in polyG and polyglutamine (polyQ) disorders. For example, glycine GGC expansions in the *NOTCH2NLC* gene cause neuronal intranuclear inclusion disease (NIID). NIID individuals with the muscle weakness phenotype carried multiple glycine GGA synonymous interruptions, whereas NIID individuals with the parkinsonism-dominant phenotype carried fewer GGA motifs and more AGC (serine) missense interruptions^16^. Additionally, intermediate-sized glutamine CAG expansions in *ATXN2* that specifically confer risk for Parkinson’s disease, Lewy body dementia and ALS, contain glutamine GAA synonymous interruptions that alter repeat motif composition without changing the encoded amino acid^17,18^. Interruptions in repeat sequences are primarily recognised for their protective role in maintaining genomic stability by preventing slippage during DNA replication and repair^19^. Beyond this canonical function, interruptions may also modulate variability in disease phenotype and clinical presentation, although the exact mechanism driving these effects remain unclear. This highlights the importance of assessing motif composition and configuration when examining association of STR expansions with disease. Given the phenotypic overlap and genetic pleiotropy between SCA and ALS and increasing awareness of the importance of motif compositions in STRs, we aimed to evaluate whether *ZFHX3* GGC repeat expansions are associated with ALS and characterise the motif composition and configuration in a large ALS and control cohort of European ancestry.

## Methods

### Standard protocol approvals, registrations and patient consents

This study was approved by the human research ethics committees of Macquarie University (ID: 10134) and by the institutional review boards of all participating centres within Project MinE. Written informed consent was provided from all participants for investigation and publication at their respective centres and additional approval was obtained from the Medical Ethical Testing Committee NedMec and the Biobanks Testing committee of UMC Utrecht. Patients were included irrespective of their carrier status for variants in known ALS-associated genes.

### International ALS cohort and control participants

A total of 6,114 people with ALS and 8,030 healthy controls were initially included in this study. ALS was diagnosed by a neurologist according to the El Escorial criteria or based on progressive weakness in the upper and lower motor neurons, confirmed by electrophysiology of widespread muscle denervation and no other diagnosis being found on neuroimaging^20^. The ALS cohort comprised 943 Australian participants recruited from the Macquarie University Neurodegenerative Disease Biobank and the Australian MND DNA Bank. The remaining 5,171 individuals with ALS were obtained from Project MinE data freeze 2; an international ALS genomics consortium spanning 14 countries^21^ summarised in Supplementary Table 1. All individuals with ALS were genotyped for the hexanucleotide repeat expansion in *C9orf72* and known pathogenic ALS variants in *SOD1, TARDBP*, and *FUS*. The control cohort comprised of 1,801 healthy individuals included in Project MinE data freeze 2 and 6,229 controls from the gnomAD European (non-Finnish) short tandem repeat data download v3.1.2.

To retain individuals of European ancestry for this study, ALS genotype call sets for all ALS cases and Project MinE controls were merged with the combined HapMap phase II and III datasets consisting of 11 populations^22^. Principal component analysis was performed using PLINK V1.9^23^ to identify samples clustering with the HapMap CEU and TSI populations, representing European ancestry. Duplicate samples were detected and removed using PLINK v1.9. A single proband was selected from ALS families where multiple individuals were available for analysis.

### *ZFHX3* GGC repeat expansion detection, validation and motif composition

Whole genome sequencing (WGS) data were available for all ALS cases and Project MinE controls of European ancestry. Details on WGS sequencing and data processing are described in the eMethods. The *ZFHX3* GGC repeat expansion was genotyped from GRCh38 aligned BAMS using ExpansionHunter^24^ v5 with the gnomAD variant catalogue excluding off target regions (json provided in eMethods). Individual-level ExpansionHunter outputs were merged into a cohort callset and summarised using TRTools^25^ mergeSTR and statSTR. For each allele a quality score (Q score) was calculated and then averaged for an individual level Q score, using the ExpansionHunter repeat length and confidence intervals, to align with the Q score metric reported in the gnomAD callset^26,27^. The *ZFHX3* GGC ExpansionHunter callset derived from WGS data was then merged with the gnomAD control callset. Individuals with a Q score below 0.2 were removed, based on a threshold to optimise sensitivity and specificity^26^.

73 alleles (36 cases; 37 controls) with a repeat expansion length ≥24 GGC units were visually validated using REViewer^28^ v2. 12 alleles (4 cases; 9 controls) were incorrectly estimated by three or more repeats upon visualisation in REViewer, as these genotypes could not be reliably corrected, they were removed from the analysis. The repeat motif composition of all 802 Australian ALS participants and a random sample of 800 healthy control participants was catalogued by manual visualisation of individual REViewer images. RStudio v5.1 was used to generate repeat length distributions and motif composition figures.

### Statistical analysis of patient clinical data and association testing

Patient phenotypic features were extracted from clinical records and variability was examined across sex, site of disease onset (bulbar or spinal), age at disease onset and disease duration (from onset until death or last known date alive). All statistical analysis was performed in RStudio v5.1. Associations between sex and site of onset were assessed using a chi-squared test of independence. Welch’s two sample t tests were used to evaluate differences in age at disease onset by both sex and site of onset. Kaplan-Meier survival analysis compared disease duration across both sex and site of onset. A linear regression model was fitted to assess the relationship between age at onset and duration for deceased patients only. Alleles with repeats lengths greater than 21 units (the median repeat length) were retained to perform a receiver operating characteristic (ROC) curve analysis and to calculate the Youden’s J statistic using the R package, cutpointr^29^. Fisher’s exact tests were used to evaluate the association of repeats expanded ≥24 GGC units and the presence of pure polyG compositions with disease status.

### Data availability

Individual-level *ZFHX3* repeat allele size data for 5,785 people with ALS and 7,982 healthy controls plus individual-level motif composition data for the subset of 802 Australian cases with ALS and 800 healthy controls is available at Zenodo (DOI: 10.5281/zenodo.18931240).

Code written in R is available in R Markdown workbooks in a GitLab repository: https://gitlab.com/mq-mnd/grp_williams/zfhx3_analysis_publication_2026.

### Results

#### Cohort summary

After filtering the cohort for ancestry and repeat quality, 5,785 participants with ALS (802 Australian ALS; 4,983 Project MinE ALS) and 7,982 healthy controls (6,215 gnomAD controls; 1,767 Project MinE controls) were retained for downstream analysis. Cohort demographic and clinical features of 13,767 individuals are summarised in Table 1. The ALS cohort comprised 61% males while 74% of controls were male. Of 5,785 people with ALS, 430 carried a known ALS disease-causal variant (Supplementary Table 2).

**Table 1.** Demographic and clinical information for study cohort.

|  | **ALS**<br>(*n* = 5,785) | **Controls**<br>(*n* = 7,982) |
| --- | --- | --- |
| Sex (Males) | 3,504 (61%) | 4,635 (74%) |
| Age at disease onset (years) | 59.7 ± 12.7 | - |
| Disease duration (months) | 9.80 ± 21.1 | - |
| **Site of onset** |  |  |
| Bulbar | 1,490 (27%) | - |
| Spinal | 3,836 (68%) | - |
| Age at disease onset and disease duration are presented as mean ± standard deviation. Age at onset was available for 5,563/5,785 individuals, disease duration was available for 5,445/5,785 individuals and site of onset was available for 5,612/5,785 individuals with ALS. |  |  |

Differences in site of onset (*P* = 2.3e-27) and age of onset (*P* = 3.8e-13) between male and females were observed. 71% of males presented with spinal onset versus 59% of females (Figure 1A). The average disease onset was 61.2 years in females and 58.7 in males (Fig 1B). Males presented with a longer disease duration than females (Fig 1C; *P* = 1.9e-06). Individuals who presented with ALS at a younger age had a longer disease duration (Fig 1D; *P* = 1.06e-11). 66% of people with ALS had spinal onset. People with spinal onset had a younger age of onset (*P* = 2.1e-47; Fig. 1E) and slower disease progression (*P* = 3.4e-31) compared to bulbar onset cases (Fig 1E and F). This is concordant with existing published ALS cohorts^6, 30^.

**Figure 1.**
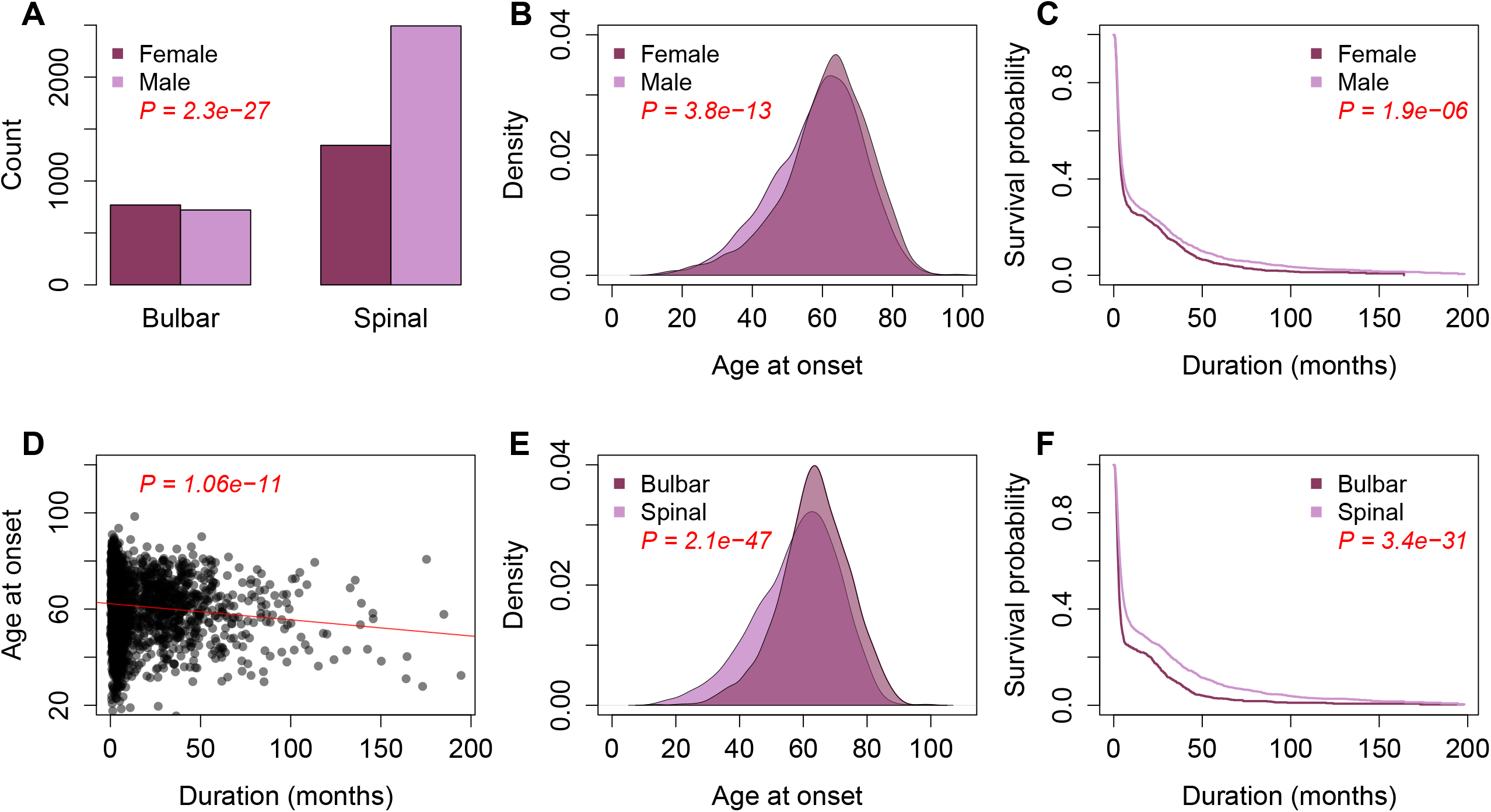
Statistical analysis of clinical variables in the ALS cohort (n = 5,785) Significant associations are denoted by red *P* values. (**A)** There was a significant difference in site of onset between males and females with ALS, as more males presented with spinal onset. **(B)** Age at disease onset was significantly older in females with ALS. **(C)** Males with ALS have a significantly longer disease duration than females. **(D)** Individuals diagnosed with ALS at a younger age lived significantly longer than those diagnosed later in life. **(E)** People with bulbar onset ALS presented with disease at an older age. **(F)** People with bulbar onset experienced a significantly shorter disease duration than people with spinal onset.

#### Frequency of *ZFHX3* GGC repeat lengths

The distribution of *ZFHX3* GGC repeat lengths across 11,270 ALS alleles and 15,964 healthy control alleles was visualised (Figure. 2). Repeat sizes ranged from 4-30 units in people with ALS and 5-29 in healthy controls. No individuals exceeded the pathogenic threshold of 42 GGC repeat units reported in SCA4 families (Supplementary Table 3). ROC analysis and Youden’s J statistic indicated that repeat length 24 was the optimal threshold to test for an association between cases and controls (Supplementary Figure 1). Yet a Fisher’s exact test revealed no association between expanded GGC repeats ≥24 in people with ALS compared to healthy controls *(P* = 0.088).

**Figure 2.**
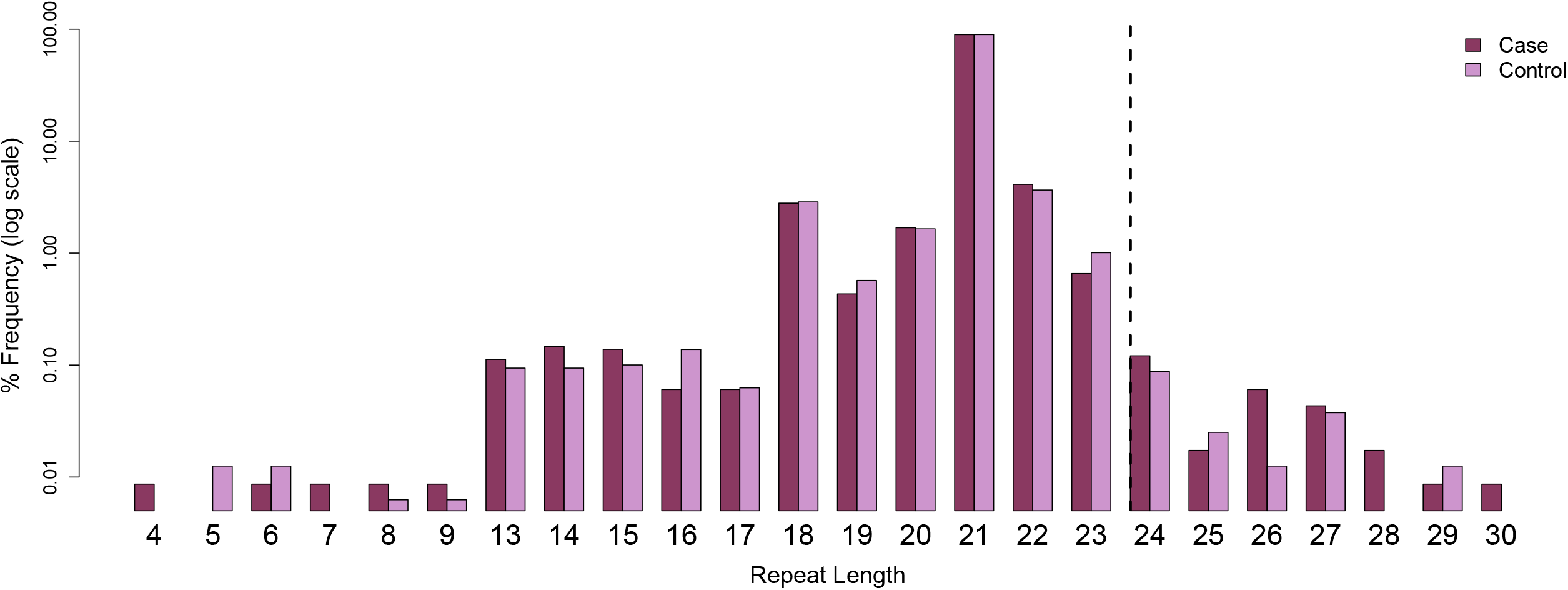
Frequency of *ZFHX3* GGC repeat lengths in ALS cases and controls. ROC analysis and Youden’s J statistic identified repeat length 24 as the optimal threshold for association test, indicated by the dashed line. However, the frequency of expanded *ZFHX3* GGC repeats ≥24 in ALS cases was not significantly increased compared to controls. The y-axis is a log scale of percent frequency to allow better depiction of frequency variations.

#### Variability of repeat motif compositions and configurations in *ZFHX3*

The frequency of 50 distinct repeat motif compositions from 802 Australian ALS individuals and 800 healthy controls across both alleles was visualised (Figure 3). All repeat motif compositions were interrupted by at least one GGT (glycine) motif that produces a synonymous interruption in the polyglycine tract. Two-thirds of these distinct motifs (32/50) were present in a single individual. Eleven motif compositions across 21 alleles (9 cases, 12 controls) lacked a missense interruption and coded pure glycine tracts, ranging in size from 5-26 repeat motifs. One individual with ALS carried a pure polyglycine tract on both alleles, with repeat lengths 23 and 26. A Fisher’s exact test revealed no association between pure polyglycine tracts in people with ALS compared to healthy controls (*P* = 0.521). The most common motif composition (in 84.7% of ALS alleles and 84.3% of control alleles) was present in alleles with 21 repeats, and comprised 18 GGC (glycine) motifs, interrupted by two GGT (glycine) motifs and one AGT (serine) motif (Figure 3 and Supplementary Figures 2 and 3). Each repeat length had multiple configurations in their motif composition, with the exception of repeat lengths 5, 25 and 26, which each had only one configuration. Alleles with a repeat length of 19 and 23 had the greatest number of different motif compositions, each with eight distinct configurations identified. Whereas alleles with a repeat length of 21 showed the highest variability in interrupting motifs, with five distinct noncanonical GGC motifs observed across six different motif compositions. Double missense interruptions occurred in 137 alleles (74 cases, 63 controls), most commonly involving combinations of AGT (serine) and GAC (aspartic acid) motifs. A single ALS allele with an AGC motif co-occurred with an AGT motif, resulting in a double serine interruption. A detailed breakdown of the motif composition seen in 1,604 ALS alleles and 1,600 control alleles is provided in Supplementary Figures 2 and 3 and Supplementary Table 4.

**Figure 3.**
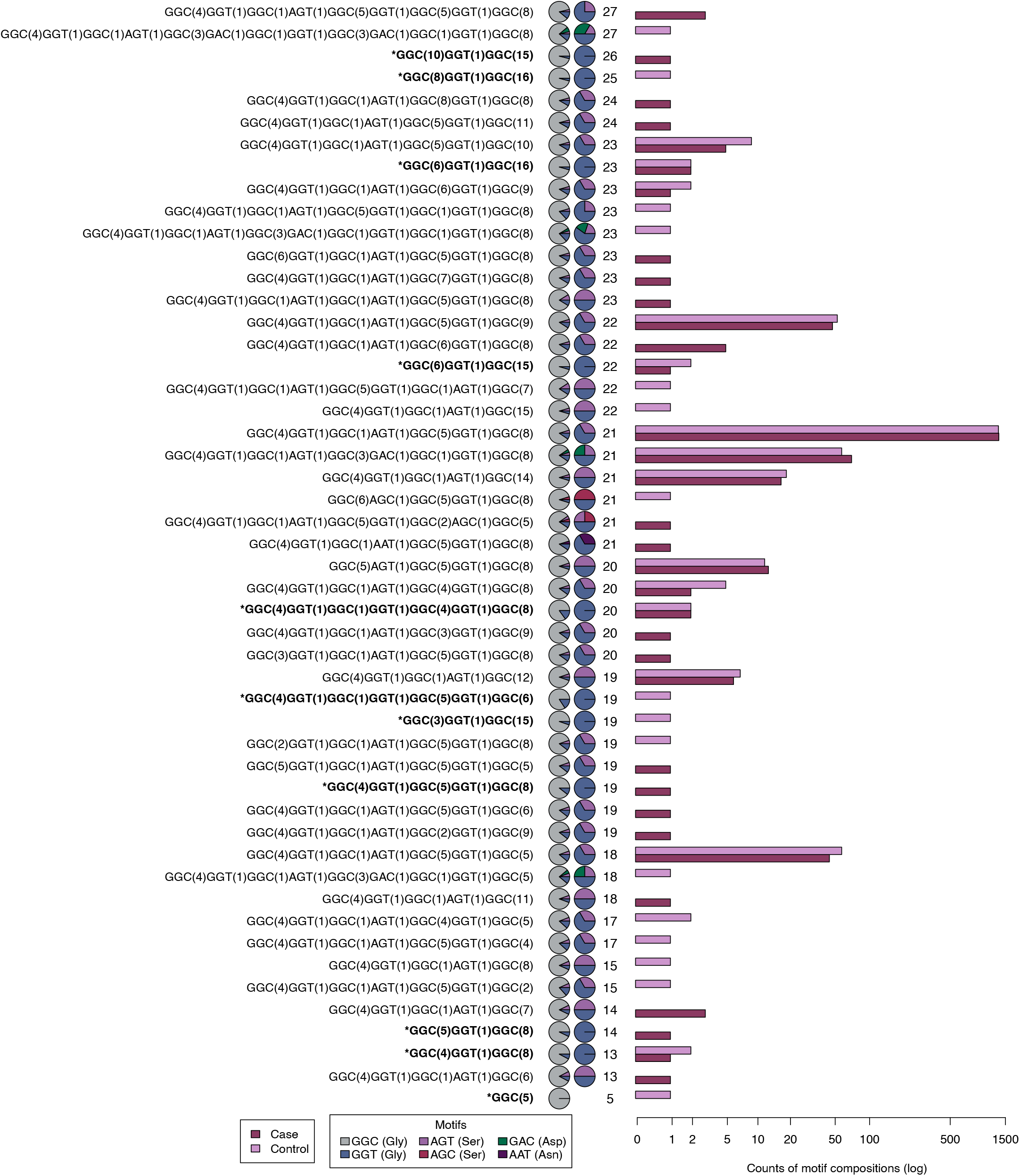
Frequency of 50 distinct repeat motif compositions in 802 Australian ALS cases and 800 controls. Frequency of distinct repeat motif compositions across 1,604 alleles from 802 Australian ALS cases and 1,600 alleles from 800 healthy controls, ordered by repeat length. For each motif composition, the first column of pie charts shows the proportional contribution of all repeat motifs, while the second column shows motif proportions after excluding the canonical GGC repeat. An asterisk (*) denotes a repeat motif composition lacking a missense interruption and represents a pure glycine tract. Counts are displayed on a log scale to allow for easier depiction of frequency variability.

### Discussion

A pathogenic GGC repeat expansion in *ZFHX3* was recently reported as the cause of SCA4^9,10,11^. Given that intermediate expansions at other SCA loci, including *ATXN1* and *ATXN2*, are associated with increased risk of ALS^6,7,8^, and the pathological overlap of SCA4 and ALS^10^, we investigated whether *ZFHX3* GGC expansions similarly contribute to ALS risk. Using ExpansionHunter-derived genotypes from whole-genome sequencing data, we found no evidence of association between expanded *ZFHX3* GGC repeats and ALS in a large European cohort, indicating that repeat length does not contribute to the risk of ALS.

Beyond repeat size, motif composition and sequence architecture are increasingly recognised as determinants of pathogenicity in repeat expansion disorders. In polyglutamine (polyQ) and polyglycine (polyG) diseases, interruption pattern, motif order, and tract purity can influence repeat instability, penetrance, and age at disease onset^14,16, 31, 32^. In our cohort, we identified 50 distinct *ZFHX3* repeat motif configurations among 802 individuals with ALS and 800 healthy controls. Two-thirds of these distinct configurations were present in only one individual. All contained at least one synonymous or missense interruption, alongside eleven compositions that encode uninterrupted polyG tracts. In contrast, pathogenic SCA4 expansions consist of long, pure, uninterrupted polyG tracts^9, 10^. Pure repeat tracts are thought to be more prone to replication slippage and somatic instability, making them susceptible to further expansion^19^, and conversely, interrupted tracts may confer locus stability. In addition, the interruption pattern and motif configuration of repeats may influence transcription and RNA processing independent of repeat length^33^. Understanding how synonymous or missense interruptions influence and stabilise repeat expansions may inform therapeutic strategies that aim to introduce interruptions to stabilise pathogenic repeat tracts, a strategy currently being explored in other repeat expansion disorders^34^. Together, our findings reinforce the importance of resolving repeat motif configuration, not solely allele size, when assessing the pathogenicity of expanded repeats.

This study had several limitations. Although cohort size was substantial, the analysis was restricted to individuals of European ancestry. Since ALS is genetically heterogeneous with mutation frequencies that vary across ancestral backgrounds^35, 36^, future studies that incorporate diverse ancestral populations will be important to more definitively evaluate the contribution of *ZFHX3* GGC expansions and motif configurations to ALS risk. In addition, while short-read sequencing datasets are widely available for ALS genomic studies, short-read data has inherent limitations for STR analysis. Repeat lengths that exceed the read length cannot be directly spanned, potentially leading to underestimation of larger repeat sizes. GC-rich motifs can introduce coverage bias and interrupted or structurally complex repeats may not be reliably resolved with short-read data. Indeed, a combination of short- and long-read sequencing was required to initially characterise the pathogenic *ZFHX3* GGC expansion in SCA4 families^9,10,37^. However, once the locus was defined, short-read sequencing was sufficient to reidentify individuals with SCA4 and detect previously unresolved ataxia and sensory neuropathy cases harbouring the *ZFHX3* GGC expansion^9^. Moreover, the repeat length distribution of non-disease alleles was consistent with that observed here^9,37^. Although repeat length was evaluated in a large number of both cases and controls, detailed motif composition was manually characterised only in a subset of the cohort. The population frequency and broader distribution of these configurations may require systematic assessment in more comprehensive populations.

In conclusion, we found no evidence that expanded *ZFHX3* GGC repeats contribute to ALS risk, and no association between pure polyG tracts and ALS in a large European cohort. However, the marked diversity of repeat motif configurations at this locus highlights the importance of interrogating sequence composition, not solely allele size, when evaluating STRs in neurodegenerative disease. Future studies that integrate long-read sequencing, diverse populations, and detailed phenotypic data will be essential to fully define the genetic and biological relevance of *ZFHX3* repeat variation in ALS and related disorders.

## Data Availability

Individual-level *ZFHX3* repeat allele size data for 5,785 people with ALS and 7,982 healthy controls plus individual-level motif composition data for the subset of 802 Australian cases with ALS and 800 healthy controls is available at

https://doi.org/10.5281/zenodo.18931240

https://gitlab.com/mq-mnd/grp_williams/zfhx3_analysis_publication_2026

## Acknowledgements

Biospecimens and related clinical data from Australian participants were obtained from the Macquarie University Neurodegenerative Disease Biobank and the Australian MND DNA Bank. The authors would like to acknowledge Susan D’Silva, Lorel Adams, Srestha Mazumder and Rosie Fell for compiling and collating clinical and demographic data. The authors would also like to gratefully acknowledge the participants who provided biological samples for this study. The collaboration project is co-funded by the PPP Allowance made available by Health∼Holland, Top Sector Life Sciences & Health, to stimulate public-private partnerships. This study was supported by the ALS foundation Netherlands. Project MinE work was sponsored by NOW-Domain science for the use of supercomputer facilities.

## Author contributions

Z. N. Zussa: Draft/revision of manuscript for content, major role in acquisition of data, analysis or interpretation of data. A.N. Smith: major role in acquisition of data, analysis or interpretation of data. J. J.F.A Van Vugt: major role in acquisition of data, analysis or interpretation of data. D. S. O’Shaughnessy: interpretation of data. N. Grima: major role in acquisition of data. S. Chan Moi Fat: interpretation of data. I. P. Blair: major role in acquisition of data. D. B. Rowe: major role in acquisition of data. R. Pamphlett: major role in acquisition of data. G. A. Nicholson: major role in acquisition of data. M. C. Kiernan: major role in acquisition of data. W. V. Rheenen: major role in acquisition of data J. Veldink: major role in acquisition of data. K. L. Williams: Draft/revision of manuscript for content, major role in acquisition of data, analysis or interpretation of data, study concept or design, supervision. L. Henden: Draft/revision of manuscript for content, major role in acquisition of data, analysis or interpretation of data, study concept or design, supervision.

## Funding

MotorOn scholarship to Z. N. Zussa. FightMND and the National Health and Medical Research Council of Australia (GNT2033019) fellowships to K. L. Williams. Motor Neuron Disease Australia Innovator Grant to L. Henden. FightMND Angie Cunningham PhD Scholarship and Project Grant-In-Aid Award to N. Grima and I. P. Blair. National Health and Medical Research Council funding to M. C. Kiernan.

## Disclosure

DBR reports consultancies or advisory boards for Biogen, Celosia Therapeutics, and Genetic Signatures. JHV reports to have sponsored research agreements with Biogen, Eli Lilly, Trace and Astra Zeneca.

## AI statement

During the preparation of this manuscript (February 2025) the authors used ChatGPT 5.2 to improve the readability of content already written by the authors. After using this tool/service, the authors reviewed and edited the content as needed and take full responsibility of the content of the publication.

## Notes

### Author Declarations

Ethics committee of Macquarie University (ID: 10134), UMC Utrecht and participating centres within Project MinE gave ethical approval for this work.

### Summary of Updates

This version of the manuscript has been revised to document the spectrum of motifs in a control population.

