## Supplementary Information for "Characterising the motif composition and allele length distribution of *ZFHX3* GGC repeat expansions in amyotrophic lateral sclerosis"

**This PDF file includes:**

Supplementary Methods
Supplementary Figures 1–3
Supplementary Tables 1–3
Members of Project Mine Consortium
Supplementary References

### Supplementary Methods

#### Whole Genome Sequencing Data Processing

##### *Australian ALS cohort*

Genomic DNA was extracted using whole blood according to standard protocols. Samples underwent library preparation using the TruSeq PCR free library prep kit (Illumina V2.5). Prepped libraries underwent multiplex 150-bp paired-end whole genome sequencing (WGS) on the Illumina HiSeq X Ten instrument (Kinghorn Centre for Clinical Genomics, Sydney, Australia and Macrogen Oceania). Sequencing data was aligned to the GRCh38 reference genome using BWA-MEM[1]. Variant calling and joint genotyping were generated using GATKs Haplotypecaller and genotypeGVCF pipeline[2] .

##### *Project MinE Data Freeze 2*

Sequencing and data processing have been described previously[3–5]. Briefly, case and control samples were sequenced using PCR-free library preparation on the Illumina HiSeq 2000 and HiSeq X platforms to ∼35-fold coverage with 100 bp reads and ∼25-fold coverage with 150 bp reads, respectively. Sequencing data was aligned to the GRCh38 reference genome and variant calling were performed using the Illumina Isaac pipeline[6].

#### Variant Catalogue for *ZFHX3* GGC Expansion Detection using ExpansionHunter v5


```
{   "LocusId": "ZFHX3",
    "LocusStructure": "(GCC)*",
    "VariantType": "Repeat",
    "ReferenceRegion": "chr16:72787694-72787757",
    "RepeatUnit": "GCC",
    "Gene": "ZFHX3",
    "GeneRegion": "coding: polyglycine",
    "GeneId": "ENST00000268489",
    "DiscoveryMethod": "L, WGS",
    "DiscoveryYear": 2023,
    "Diseases": [
        {
            "Symbol": "SCA4",
            "Name": "Spinocerebellar ataxia 4",
            "Inheritance": "AD",
            "OMIM": "600223",
            "NormalMax": 26,
            "PathogenicMin": 41
        }
    ],
    "MainReferenceRegion": "chr16:72787694-72787757",
    "Inheritance": "AD"
}
```

### Supplementary Figures 1–3

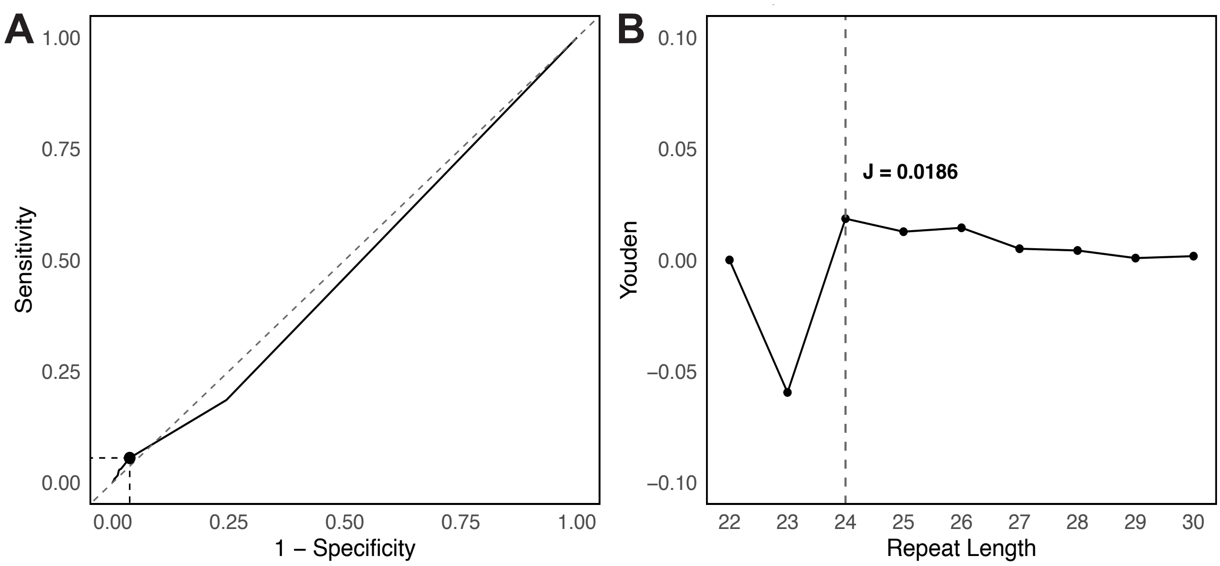


**Supplementary Figure 1: ROC analysis and Youden's J statistic of *ZFHX3* repeat expansions (22-30).** A) ROC analysis of *ZFHX3* GGC repeat expansions (22-30). Sensitivity is the true positive rate and specificity is the true negative rate. The optimal sensitivity and specificity is indicated by a black dot (n=24 repeats). B) Youden's J statistic calculated for each repeat length (22-27). The J statistic assess the sensitivity and specificity from the ROC analysis to find the optimal threshold that can discriminate cases from controls. The maximum Youden's J statistic was observed at a *ZFHX3* repeat length of 24 (J= 0.0186), indicating the optimal threshold.

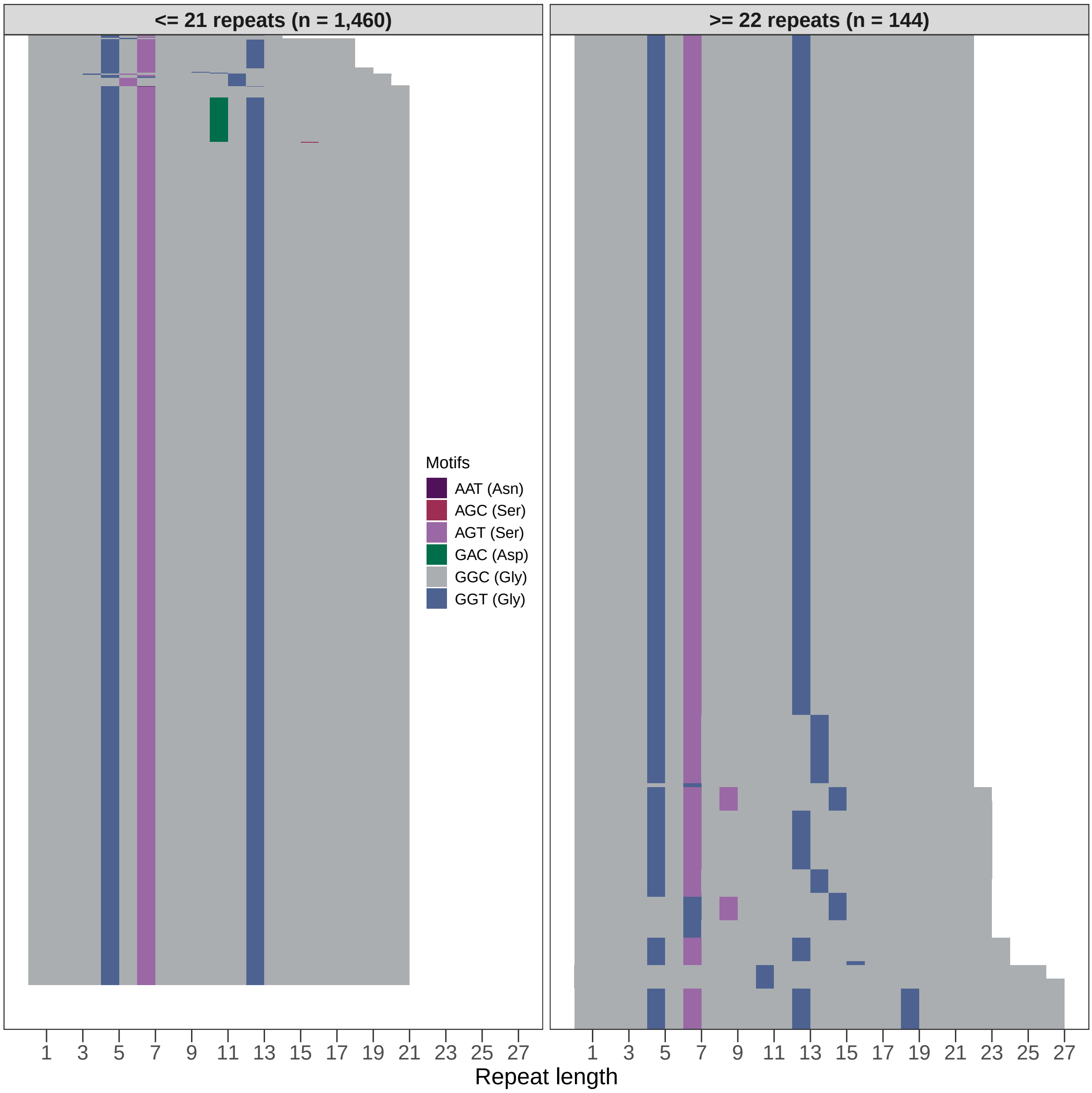


**Supplementary Figure 2: Motif composition of *ZFHX3* repeat alleles in the Australian ALS cohort (*n* = 802).** Motif composition of the long and short alleles from 802 Australian ALS patients. Alleles are ordered by increasing repeat length. The left panel shows alleles with 21 repeats or fewer. The right panel shows alleles with 22 repeats or more. Colours denote distinct repeat motifs, with grey indicating the canonical GGC repeat motif. The y-axis has been scaled according to the number of alleles with repeats either ≤21 ($n = 1,460$) or ≥21 ($n = 144$).

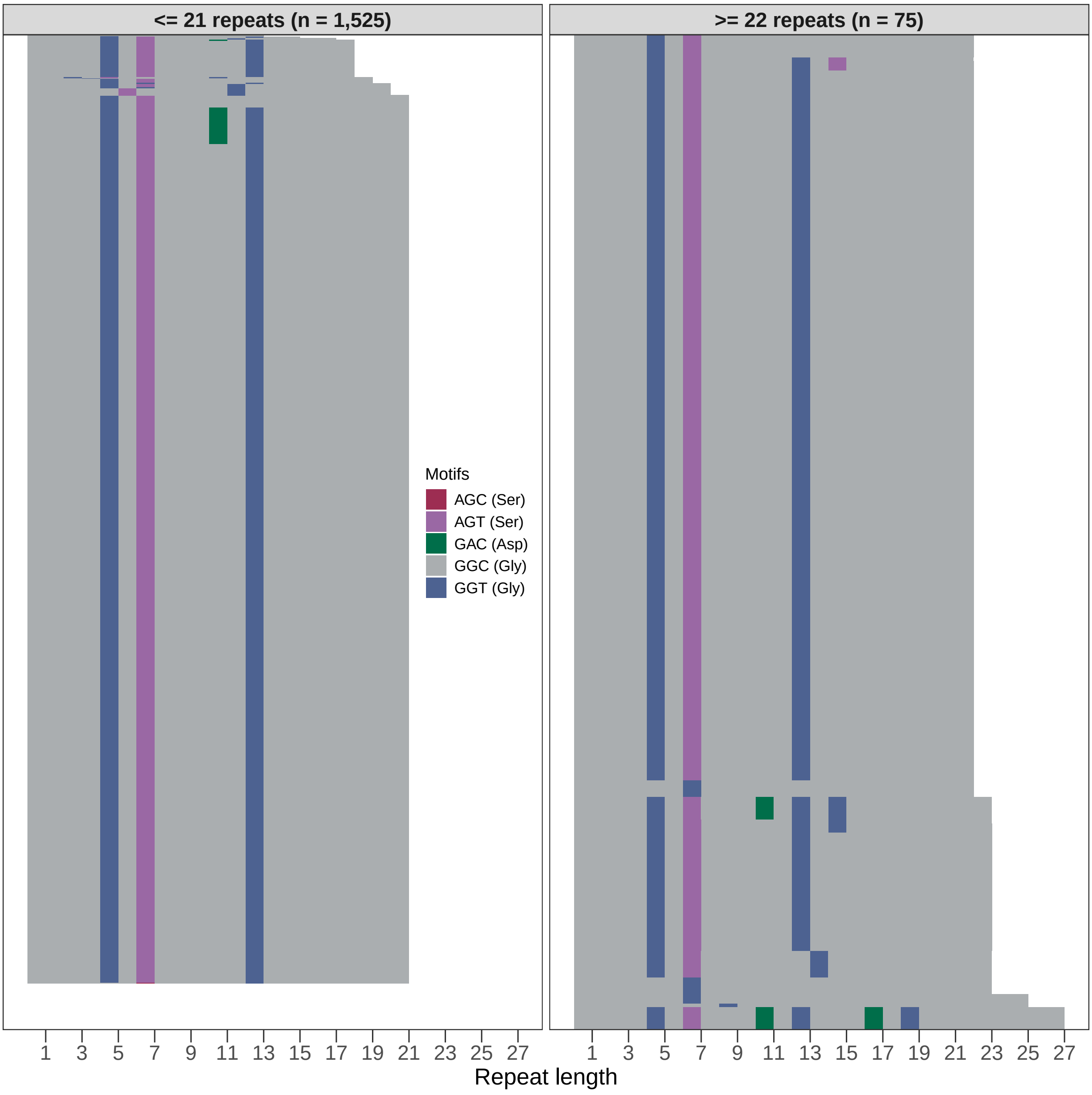


**Supplementary Figure 3: Motif composition of *ZFHX3* repeat alleles in the project MinE control cohort (*n* = 800).** Motif composition of the long and short alleles from 800 healthy controls sourced from project MinE. Alleles are ordered by increasing repeat length. The left panel shows alleles with 21 repeats or fewer. The right panel shows alleles with 22 repeats or more. Colours denote distinct repeat motifs, with grey indicating the canonical GGC repeat motif. The y-axis has been scaled according to the number of alleles with repeats either ≤21 ($n = 1,525$) or ≥21 ($n = 75$).

### Supplementary Tables 1–3

**Supplementary Table 1** Summary of participants from Project MinE DF2 and contributing countries.

| Country | ALS | Controls |
|---|---|---|
| Belgium | 552 | 183 |
| France | 246 | 39 |
| Ireland | 462 | 232 |
| Israel | 100 | - |
| Italy | 55 | - |
| Netherlands | 586 | 393 |
| Portugal | 45 | 16 |
| Spain | 370 | 165 |
| Sweden | 241 | 112 |
| Switzerland | 44 | - |
| Turkey | 608 | 141 |
| United Kingdom | 1,409 | 443 |
| United States of America | 450 | 77 |

**Supplementary Table 2** ALS-causal or -associated genetic variants present in ALS cases in this cohort[7]

| Gene | Accession number | cDNA change | Protein change | sALS cases | fALS cases |
|---|---|---|---|---|---|
| *C9orf72* | NM_001256054 | $GGGGCC^{RE}$ | | 352 | - |
| *TARDBP* | NM_007375 | c.269C>T | p.Ala90Val | 3 | - |
| *TARDBP* | NM_007375 | c.859G>A | p.Gly287Ser | 3 | - |
| *TARDBP* | NM_007375 | c.1028A>G | p.Gln343Arg | 1 | - |
| *TARDBP* | NM_007375 | c.1055A>G | p.Asn352Ser | 4 | - |
| *TARDBP* | NM_007375 | c.1144G>A | p.Ala382Thr | 1 | - |
| *TARDBP* | NM_007375 | c.1169A>G | p.Asn390Ser | 1 | - |
| *FUS* | NM_004960 | c.1292C>T | p.Pro431Leu | 3 | - |
| *FUS* | NM_004960 | c.1520G>A | p.Gly507Asp | 1 | - |
| *FUS* | NM_004960 | c.1561C>T | p.Arg521Cys | 2 | 1 |
| *FUS* | NM_004960 | c.1574C>T | p.Pro525Leu | 3 | - |
| *SOD1* | NM_000454 | c.14C>T | p.Ala5Val | 2 | - |
| *SOD1* | NM_000454 | c.25C>G | p.Leu9Val | 1 | - |
| *SOD1* | NM_000454 | c.44T>G | p.Val15Gly | 1 | - |
| *SOD1* | NM_000454 | c.59A>G | p.Asn20Ser | 2 | - |
| *SOD1* | NM_000454 | c.65A>G | p.Glu22Gly | 2 | - |
| *SOD1* | NM_000454 | c.68A>T | p.Gln23Leu | 3 | - |
| *SOD1* | NM_000454 | c.112G>A | p.Gly38Arg | 2 | - |
| *SOD1* | NM_000454 | c.197A>G | p.Asn66Ser | 1 | - |
| *SOD1* | NM_000454 | c.260A>G | p.Asn87Ser | 1 | - |
| *SOD1* | NM_000454 | c.280G>T | p.Gly94Cys | 4 | - |
| *SOD1* | NM_000454 | c.281G>C | p.Gly94Ala | 1 | - |
| *SOD1* | NM_000454 | c.302A>G | p.Glu101Gly | - | 2 |
| *SOD1* | NM_000454 | c.317C>T | p.Ser106Leu | 1 | - |
| *SOD1* | NM_000454 | c.341T>C | p.Ile114Thr | 10 | 17 |
| *SOD1* | NM_000454 | c.358G>C | p.Val120Leu | 2 | - |
| *SOD1* | NM_000454 | c.442G>A | p.Gly148Ser | 1 | - |
| *SOD1* | NM_000454 | c.446T>G | p.Val149Gly | - | 2 |

**Supplementary Table 3** Reported thresholds for *ZFHX3* GGC repeat expansions in literature.

| Resource | Normal | Intermediate | Pathogenic |
|---|---|---|---|
| gnomAD Tandem Repeats: ZFHX3[8] | $\leq 26$ | - | $\geq 41$ |
| STRchive: SCA4 ZFHX3[9] | 14–26 | 27–45 | 46–74 |
| STRipy: SCA4 ZFHX3[10] | 14–31 | - | $\geq 42$ |
| Wallenius et al., 2024[11] | 14–26 | - | 42–74 |
| Figueroa et al., 2024[12] | $\leq$21 | - | 42–74 |
| Paucar et al., 2024[13] | $\leq$26 | - | 46–64 |

#### Members of Project Mine ALS Sequencing Consortium

Philip van Damme[1], Philippe Corcia[2], Philippe Couratier[3], Patrick Vourc'h[4], Orla Hardiman[5], Russell McLaughin[6], Marc Gotkine[7], Yossef Lerner[7], Yehuda Shovman[7], Vivian Drory[8], Nicola Ticozzi[9], Vincenzo Silani[9], Jan H. Veldink[10], Leonard H. van den Berg[10], Mamede de Carvalho[11], Teresa Salas[12], Jesus S. Mora Pardina[13], Monica Povedano[14], Peter Andersen[15], Markus Weber[16], Nazli A. Başak[17], Ammar Al-Chalabi[18], Chris Shaw[18], Pamela J. Shaw[19], Karen E. Morrison[20], John E. Landers[21], Jonathan D. Glass[22], Clifton L. Dalgard[23]

[1] KU Leuven - University of Leuven, Department of Neurosciences
[2] Centre SLA, CHRU de Tours, Tours, France; UMR 1253, iBrain, Université de Tours, Inserm, Tours, France.
[3] Centre SLA CHU Dupuytren Limoges France.
[4] Service de Biochimie et Biologie moléculaire, CHU de Tours, Tours, France
[5] Academic Unit of Neurology, Trinity College Dublin, Trinity Biomedical Sciences Institute, Dublin, Republic of Ireland.
[6] Complex Trait Genomics Laboratory, Smurfit Institute of Genetics, Trinity College Dublin, Dublin, Republic of Ireland.
[7] Department of Neurology, Hadassah Medical Organization and Faculty of Medicine, Hebrew University of Jerusalem, Israel
[8] Department of Neurology Tel-Aviv Sourasky Medical Centre, Israel.
[9] Department of Neurology and Laboratory of Neuroscience, IRCCS Istituto Auxologico Italiano, Milano, Italy.
[10] Department of Neurology, UMC Utrecht Brain Center, University Medical Center Utrecht, Utrecht University, Utrecht, The Netherlands.
[11] Instituto de Fisiologia, Instituto de Medicina Molecular, Faculdade de Medicina, Universidade de Lisboa, Lisbon, Portugal
[12] ALS Unit, Hospital Universitario La Paz-Carlos III, Madrid, Spain
[13] ALS Unit, Hospital San Rafael, Madrid, Spain.
[14] la Unitat Funcional de Motoneurona, Cap de Secció de Neurofisiologia, Servei de Neurologia, Hospital Universitario de Bellvitge-IDIBELL
[15] Department of Clinical Science, Neurosciences, Umeå University, Sweden.
[16] Neuromuscular Diseases Unit/ALS Clinic, HOCH, Health Ostschweiz, Kantonsspital St. Gallen, 9007, St. Gallen, Switzerland.
[17] Koç University, School of Medicine, Molecular Biology and Genetics- KUTTAM, Suna and Inan Kıraç Foundation, Istanbul Turkey.
[18] Maurice Wohl Clinical Neuroscience Institute, King's College London, Department of Basic and Clinical Neuroscience, London, UK.
[19] Sheffield Institute for Translational Neuroscience (SITraN), University of Sheffield, Sheffield, UK.
[20] School of Medicine, Dentistry and Biomedical Sciences, Queen's University Belfast, UK.
[21] Department of Neurology, University of Massachusetts Chan Medical School, Worcester, MA, USA.
[22] Department Neurology, Emory University School of Medicine, Atlanta, GA, USA.
[23] The American Genome Center, Uniformed Services University - "America's Medical School", Bethesda, MD, USA.
